# Effect of SARS-CoV-2 infection upon male gonadal function: A single center-based study

**DOI:** 10.1101/2020.03.21.20037267

**Authors:** Ling Ma, Wen Xie, Danyang Li, Lei Shi, Yanhong Mao, Yao Xiong, Yuanzhen Zhang, Ming Zhang

## Abstract

Since SARS-CoV-2 infection was first identified in December 2019, it spread rapidly and a global pandemic of COVID-19 has occurred. ACE2, the receptor for entry into the target cells by SARS-CoV-2, was found to abundantly express in testes, including spermatogonia, Leydig and Sertoli cells. However, there is no clinical evidence about whether SARS-CoV-2 infection can affect male gonadal function so far. In this study, we compared the sex-related hormones between 81 reproductive-aged men with SARS-CoV-2 infection and 100 age-matched healthy men, and found that serum luteinizing hormone (LH) was significantly increased, but the ratio of testosterone (T) to LH and the ratio of follicle stimulating hormone (FSH) to LH were dramatically decreased in males with COVID-19. Besides, multivariable regression analysis indicated that c-reactive protein (CRP) level was significantly associated with serum T:LH ratio in COVID-19 patients. This study provides the first direct evidence about the influence of medical condition of COVID-19 on male sex hormones, alerting more attention to gonadal function evaluation among patients recovered from SARS-CoV-2 infection, especially the reproductive-aged men.

## Introduction

Since the first report in Wuhan in December 2019, a novel coronavirus-induced pneumonia (called COVID-19 by WHO) spread rapidly and triggered a global pandemic outbreak ^[1]^. COVID-19 is caused by a previously unknown beta-coronavirus which is now named SARS-CoV-2 due to its high sequence similarity (∼80%) with SARS-CoV ^[2]^. Except for the respiratory symptoms such as cough, fever and even acute respiratory failure, evidences of SARS-CoV-2 attack to multiple organs such as digestive, cardiovascular, urinary systems have been reported ^[3-5]^. Angiotensin-converting enzyme 2 (ACE2) is considered as the receptor for binding and entry into host cells by SARS-CoV-2 ^[6-7]^. Theoretically, any cells expressing ACE2 may be susceptible to SARS-CoV-2 infection. According to the online database *The Human Protein Atlas portal*, testes shows the highest expression level of ACE2 protein and mRNA in the body ^[8]^. Based on scRNA-seq profiling of human testes, Wang ZP et al. also reported that ACE2 is predominantly enriched in spermatogonia, Leydig and Sertoli cells ^[9]^. All the findings suggest the potential risk of male gonad to be vulnerable to SARS-CoV-2 attack.

In the condition of viremia, virus may seed into the male reproductive track because the blood-testes barrier is not perfect enough to completely isolate virus ^[10]^. A wide breath of viruses, such as Zika, Ebola, Marburg viruses, etc. have been found in male testes and semen ^[11]^. Virus-induced testes damage can impair gonadal hormone secretion and spermatogenesis, as seen in HIV or mumps-induced orchitis ^[12]^. Previous study on SARS suggested the SARS-CoV can cause orchitis ^[13]^. However, there is no clinical information about whether SARS-CoV-2 infection can affect male gonadal function so far. In this study, we compared the sex-related hormones between reproductive-aged men with SARS-CoV-2 infection and age-matched healthy men, and found serum luteinizing hormone (LH) was significantly increased, but the ratio of testosterone (T) to LH and the ratio of follicle stimulating hormone (FSH) to LH were dramatically decreased in male with COVID-19. This study provides the first direct evidence about the influence of medical condition of COVID-19 on male sex hormones, alerting more attention to gonadal function evaluation among patients recovered from SARS-CoV-2 infection, especially the reproductive-aged men.

## Methods

### Study design and patients

We performed a retrospective study involving 81 male patients with COVID-19 as the study group, who were hospitalized in Wuhan Leishenshan Hospital from Mar 5 to Mar 18, 2020. All cases were laboratory-confirmed as SARS-CoV-2 positive using quantitative RT-PCR (qRT-PCR) on nasal and pharyngeal swab specimens. The diagnosis of COVID-19 and the severity was determined according to the New Coronavirus Pneumonia Prevention and Control Program (7th edition) published by the National Health Commission of China. Briefly, the mild type was defined as “having malaise only without positive chest radiologic findings”; the common type was defined as “having common respiratory infection symptoms such as fever, cough and positive chest radiologic changes”; the severe type was determined if any of the following conditions existed, including dyspnea (respiratory rate ≥ 30 per minute), low finger oxygen saturation (≤93% at rest), low PaO2/FiO2 (≤ 300mmHg) or rapid progress of chest radiological abnormality (>50% within 24-48 hours); and the very severe type was determined if respiratory failure (mechanical ventilation needed), shock or multiple organ dysfunctions was complicated.

All patients aged from 20∼54 years (with a median of 38 yrs). After finishing laboratory tests required for the routine medical purposes, the residual serum samples were collected for male hormone profiles detection. The control group came from the population who previously received reproductive function evaluation and were classified as having normal fertility. 100 age-matched healthy men were randomly selected and the data of their sex-related hormones were collected.

This study was reviewed and approved by the Medical Ethical Committee of Zhongnan Hospital of Wuhan University (approval number 2020033). The residual serum samples used in this study were usually discarded as a medical waste otherwise. And we didn’t directly contact with the patients and exert no burden or harm on them. Therefore, written informed consent was waived.

### Sex-related hormone assessment

In the study group, serum testosterone (T), estradiol (E2), progesterone (P), prolactin (PRL), luteinizing hormone (LH), follicle stimulating hormone (FSH), anti-mullerian hormone (AMH) were detected by electrochemiluminescent immunoassays according to the instructions from the manufacturer (cobas e411, Roche, Switzerland). In the control group, the data of serum T, E2, PRL, FSH and LH levels were retrieved from the dataset already kept in our reproductive medical center. The ratio of T:LH, T: E2 and FSH:LH were also calculated.

### Statistical analysis

All statistical analysis was performed using Graphpad Prism 6.04 (San Diego, USA) and SPSS 16.0 (Chicago, IL, USA). Continuous variables were expressed as means ± standard deviations (SD) or medians and interquartile ranges (IQR) as appropriate. Categorical variables were summarized as the counts and percentages (%). The distribution of data was analyzed by Kologorov-Smirov test. Differences between two groups were analyzed by Student’s t test (parametric) or Mann-Whitney U test (non-parametric). Univariable and multivariable linear regression were performed to analyze the relationship between serum T:LH ratio and the clinical characteristics of the COVID-19 patients. Only the variables showing statistical significance in univariable analysis were included in the multivariable analysis. Statistical significance was defined as *p* values of < 0.05.

## Results

Among 81 men with COVID-19, 86.42% (70/81) were diagnosed as “common type”, 8.64% (7/81) as “severe type” and 2.47 % (2/81) as “very severe type”. The usages of corticosterone, arbidol, oseltamivir and intravenous antibiotics was 14.81% (12/81), 44.44 % (36/81), 33.33 % (27/81) and 51.85% (42/81) respectively. 38.27% (31/81) of the patients had elevated serum alanine transaminase (ALT) and/or serum aspartate transaminase (AST), indicating the impaired liver function. The clinical characteristics of the patients were presented in Table 1.

**Table 1.**
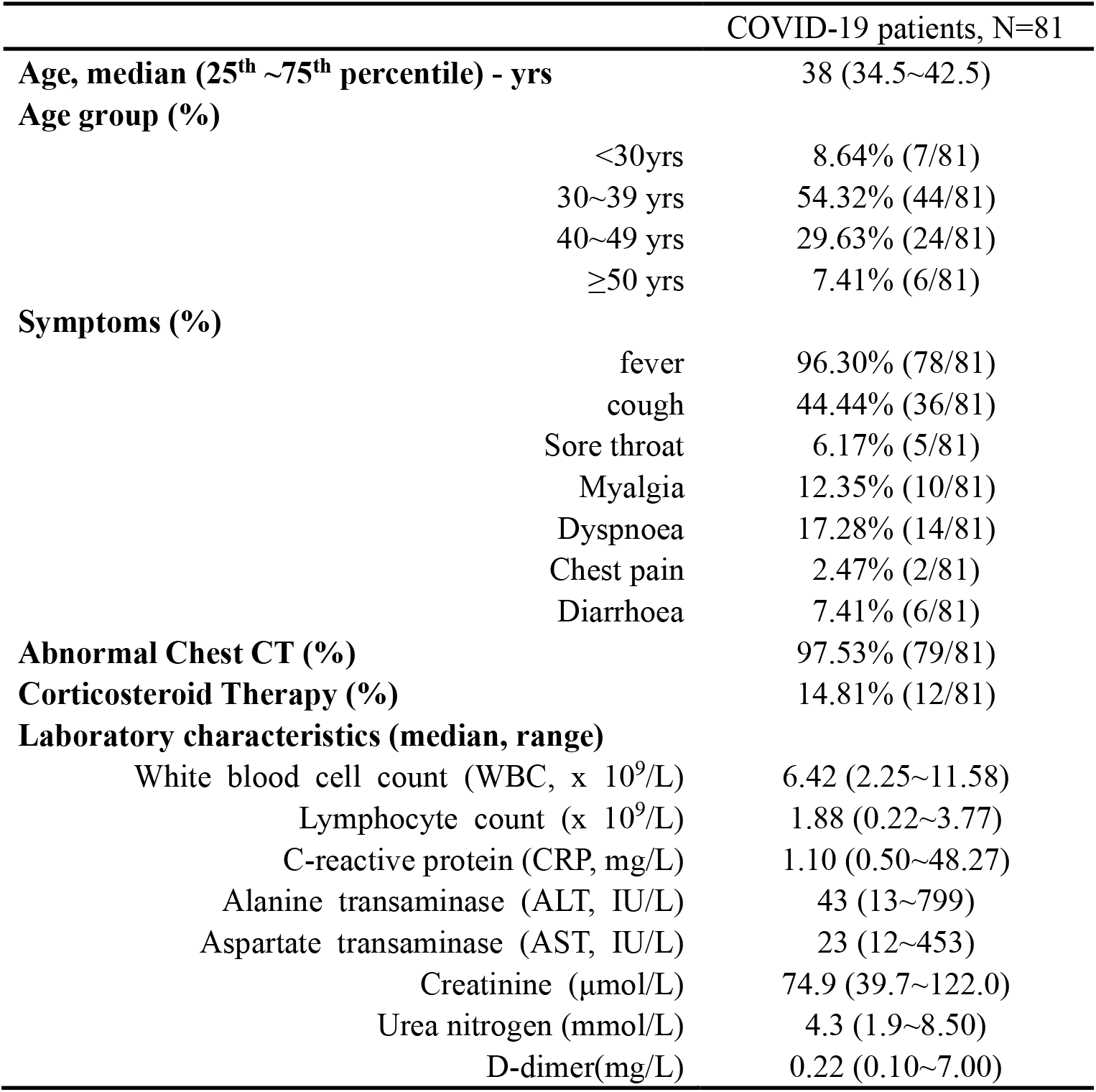
Clinical characteristics of 81 male patients with COVID-19

Compared to the control group, COVID-19 patients had significantly higher serum LH (p<0.0001) and serum PRL (p<0.0001). Although there was no statistical difference in serum T (p=0.0945) or FSH (p=0.5783) between the two groups, the ratios of T:LH (p<0.0001) and FSH: LH (p<0.0001) were dramatically decreased in the COVID-19 group (seen in Table 2).

**Table 2.** Sex-related Hormone Profiles in COVID-19 group and the control group

|  | Men with COVID-19<br>(N=81) | Age-matched healthy men<br>(N=100) | *p* value |
| --- | --- | --- | --- |
| Age - yrs | 38 (34.5~42.5) | 38 (35~41) | 0.7021 |
| Testosterone (ng/mL) | 3.97(3.12~5.68) | 4.79 (3.49~5.62) | 0.0945 |
| FSH (mIU/mL) | 4.13(3.02~6.31) | 4.51 (3.39~5.97) | 0.5783 |
| LH (mIU/mL) | 5.93(4.31~8.25) | 3.28 (2.48~4.61) | <0.0001* |
| PRL (ng/mL) | 24.07(18.76~31.15) | 7.82(5.92~11.22) | <0.0001* |
| T / LH | 0.74 (0.44~1.06) | 1.31(0.95~2.06) | <0.0001* |
| FSH / LH | 0.79 (0.51~1.01) | 1.38 (1.00~1.91) | <0.0001* |
| AMH (ng/mL) | 6.20(4.21~9.59) | N/A | N/A |

Data are presented as medians (25<sup>th</sup> ~75<sup>th</sup> percentile).
\* statistically significant

In the control group, only 54 out of 100 men had the record of serum E2. Since they were also age-matched with the COVID-19 patients, the comparison of E2 or T: E2 ratio was performed between them and the COVID-19 group. No significant difference was observed in either E2 or T: E2 ratio. (seen in Table 3)

**Table 3.** Serum Estradiol (E2) and T: E2 ratio in COVID-19 paitents and the age-matched healthy men

|  | Men with COVID-19<br>(N=81) | Age-matched healthy men*<br>(N=54) | *p* value |
| --- | --- | --- | --- |
| Age (year) | 38.00 (34.50~42.50) | 38.5 (36.0~44.25) | 0.6896 |
| Estradiol (pg/mL) | 32.34 (27.06~41.55) | 33.00 (23.75~38.00) | 0.3183 |
| T / E2 | 0.12 (0.10~0.15) | 0.14 (0.10~0.18) | 0.1340 |

\* Only 54 out of 100 in the control group had the record of serum estradiol. Since they are also age-matched with the COVID-19 group, they are used as the control group for E2 and T: E2 comparison.
Data are presented as medians (25<sup>th</sup> ~75<sup>th</sup> percentile).

By univariable linear regression analysis, it can be seen that serum T:LH ratio in the COVID-19 group was negatively associated with the severity (p=0.0236), aspartate transaminase (AST) concentration (p=0.0287), and c-reactive protein (CRP) level (p<0.0001), but positively associated with serum AMH level (p=0.0067). On multivariable analysis, only CRP level (p=0.0128) were significantly associated serum T:LH. (Seen in Table 4)

**Table 4.** Linear regression for association of clinical characteristics of COVID-19 patients with serum T:LH ratio

|  | Univariable analysis |  |  | Multivariable analysis |  |  |
| --- | --- | --- | --- | --- | --- | --- |
|  | 95% CI (slope) |  | *p* value | 95% CI |  | *p* value |
| Severity † | -0.4564 | -0.0338 | 0.0236 | -0.2515 | 0.1719 | 0.7083 |
| AST (IU/L) | -63.65 | -3.593 | 0.0287 | -0.0017 | 0.0014 | 0.8864 |
| CRP (mg/L) | -15.71 | -6.020 | <0.0001 | -0.0281 | -0.0035 | 0.0128* |
| AMH (ng/mL) | 0.7733 | 4.656 | 0.0067 | -0.0093 | 0.0393 | 0.2233 |

Only the variables showing statistical significance in univariable analysis were included in the multivariable analysis.
95% CI: 95% confidence interval
\* statistically significant
† severity: 1- mild type, 2-moderate type, 3-severe type, 4-critical type

## Discussion

There are accumulative evidences that male reproductive systems are vulnerable to virus infection. Unlike bacterial infections which usually invade accessory glands and epididymis, virus circulating in the blood mainly attack testis. It is known that a broad range of virus families, including human immunodeficiency virus (HIV), mumps virus, influenza, Zika virus, CoxsacKie virus, may induce orchitis and even result in male infertility ^[14]^. Besides, many viruses such as Ebola, HIV, Zika and Hepatitis viruses B/C can be transmitted into semen and cause sexual transmission. The deleterious effects of viruses involve the direct damage of spermatozoon, abnormal sex-hormone secretion, and dysregulation of inflammatory cytokines. For example, a study based on rams showed that bluetongue virus can replicate in the endothelial cells of the peritubular areas within the testes, leading to enhanced type-I interferon response, decreased testosterone biosynthesis by Leydig cells, and even destruction of Sertoli cells ^[15]^.

The testes are mainly constructed by seminiferous tubules and intertubular tissue. The seminiferous tubules are the place where the sperm are generated, composed of sperm-producing cells (spermatogonia) and the supporting Sertoli cells. The interstitial Leydig cells are responsible for testosterone production under regulation of LH. It was reported that SARS-CoV-2 uses ACE2 and the cellular serine protease TMPRSS2 for entry into host cells ^[6]^. Based on scRNA-seq analysis, Wang ZP et al. found that TMPRSS2 mainly exists in spermatogonia and spermatids, whereas ACE2 widely expresses in spermatogonia, Leydig and Sertoli cells. Gene ontology (GO) enrichment analysis further indicated that GO categories associated with viral reproduction and transmission were positively enriched in ACE2-positive spermatogonia ^[9]^. SARS, the similar virus of SARS-CoV-2, has been reported to cause orchitis ^[13]^. Taken together, we suppose that testes may also run high risk of damage and dysregulation under COVID-19.

Since the major roles of testes are spermatogenesis and androgens secretion, the sex-related steroids can be used to evaluate the status of male gonad. In order to learn the effect of SARS-CoV-2 infection on male reproductive function, we compared the sex hormone profiles between COVID-19 patients and age-matched healthy men with normal fertility in this study. Although serum testosterone levels did not statistically change in the COVID-19 group, a significant increase in serum LH level and a dramatic decrease in serum T: LH were observed. In interpreting those results, the following points should be taken into account: (1) As known, there is a subtle negative feedback between T in testes and LH in pituitary. In the early stage of hypogonadism, impaired T production may stimulate the release of LH which can maintain T level temporarily. (2) The basal T level in the population varies widely, thus the ratio between hormones, such as T/LH or T/E2, was considered as better parameters for male gonad function evaluation ^[16-17]^.

The serum PRL level also significantly elevated in COVID-19 patients. Since serum PRL can be influenced be multiple factors, such as diet, stress, drugs, etc., the elevation was not surprising. But it should be mentioned that high PRL level may lead to pituitary suppression and decreased gonadotropins ^[18]^, while serum LH was increased in this study. Taken together, we infer that the elevated LH and decreased T:LH ratio are more likely to be caused by testes dysfunction, such as the possible damage of Leydig cells.

Unlike serum LH level, serum FSH, serum E2 and the ratio of T: E2 were not significantly different between the COVID-19 group and the control group. In men, FSH is mainly suppressed by inhibin B secreted by Sertoli cells, and estradiol normally comes from peripheral aromatization of androgens. Therefore, it seemed that Sertoli cells were less disturbed than Leydig cells under COVID-19.

We also analyzed the relationship between serum T:LH and main clinical characteristics of the COVID-19 patients. Although higher rank of severity, elevated AST, increased CRP and AMH seemed to be associated with lower T:LH on univariable analysis, only CRP was significantly associated with T:LH ratio after adjustment on multivariable analysis. CRP is an acute-phase protein produced by the liver which rises in acute inflammation throughout the body. In COVID-19, rapid and dramatic increase of CRP was observed more often in severe cases than in non-severe cases ^[19]^. In acute inflammation, elevated CRP are also accompanied with abnormal cytokines. And some cytokines, such as interferon, may affect testes function and spermatogenesis ^[20]^.

This study has several strengths. The study provides the first evidence about the alteration of sex-related hormones under COVID-19. And we found serum LH significantly elevated and T: LH ratio decreased in COVID-19 patients, which infer to the potential hypogonadism. The percentage of non-severe cases (mild and common type) included in this study is 88% (72/81), which is consistent to what has been reported by a large retrospective study ^[20]^. Since more than half of people with COVID-19 were reproductive-aged ^[20]^, more attention should be paid to the effect of SARS-CoV-2 on reproductive system, and gonadal function evaluation including semen examination is necessary in the follow-up of those who recovered from COVID-19.

There are also some limitations in this study. First, neither semen parameters nor existence of SARS-CoV-2 in semen was detected, which are more straightforward evidence for testes injury caused by SARS-CoV-2. Second, due to the small size of samples, only 2 mild cases, 7 severe cases and 2 very severe cased are included in, which may affect the power of statistical analysis, such as the association between severity and T:LH ratio. Third, in the condition of COVID-19, some other factors such as stress and corticosteroid therapy may also influence hypothalamic-pituitary-gonadal axis. However, it should be mentioned that corticosteroids are usually believed to impair LH release instead of promoting it as seen in this study ^[21,22]^. In the end, repeated detection with appropriate time interval (such as 3 months or 6 months later) is necessary.

## Data Availability

The data used to support the findings of this study are available from the corresponding author upon request

